# A Nanopore sequencing-based pharmacogenomic panel to personalize tuberculosis drug dosing

**DOI:** 10.1101/2023.09.08.23295248

**Authors:** Renu Verma, Kesia Esther da Silva, Neesha Rockwood, Roeland E. Wasmann, Nombuso Yende, Taeksun Song, Eugene Kim, Paolo Denti, Robert J. Wilkinson, Jason R. Andrews

## Abstract

**Rationale:** Standardized dosing of anti-tubercular (TB) drugs leads to variable plasma drug levels, which are associated with adverse drug reactions, delayed treatment response, and relapse. Mutations in genes affecting drug metabolism explain considerable interindividual pharmacokinetic variability; however, pharmacogenomic (PGx) assays that predict metabolism of anti-TB drugs have been lacking.

**Objectives:** To develop a Nanopore sequencing panel and validate its performance in active TB patients to personalize treatment dosing.

**Measurements and Main Results:** We developed a Nanopore sequencing panel targeting 15 single nucleotide polymorphisms (SNP) in 5 genes affecting the metabolism of isoniazid (INH), rifampin (RIF), linezolid and bedaquiline. For validation, we sequenced DNA samples (n=48) from the 1000 genomes project and compared variant calling accuracy with Illumina genome sequencing. We then sequenced DNA samples from patients with active TB (n=100) from South Africa on a MinION Mk1C and evaluated the relationship between genotypes and pharmacokinetic parameters for INH and RIF.

**Results:** The PGx panel achieved 100% concordance with Illumina sequencing in variant identification for the samples from the 1000 Genomes Project. In the clinical cohort, coverage was >100x for 1498/1500 (99.8%) amplicons across the 100 samples. One third (33%) of participants were identified as slow, 47% were intermediate and 20% were rapid isoniazid acetylators. Isoniazid clearance was significantly impacted by acetylator status (p<0.0001) with median (IQR) clearances of 11.2 L/h (9.3-13.4), 27.2 L/h (22.0-31.7), and 45.1 L/h (34.1-51.1) in slow, intermediate, and rapid acetylators. Rifampin clearance was 17.3% (2.50-29.9) lower in individuals with homozygous *AADAC* rs1803155 G>A substitutions (p=0.0015).

**Conclusion:** Targeted sequencing can enable detection of polymorphisms influencing TB drug metabolism on a low-cost, portable instrument to personalize dosing for TB treatment or prevention.

**Summary:** This manuscript describes the development and validation of Nanopore sequencing panel to detect host pharmacogenomic markers to guide personalized drug dosing for treatment or prevention of tuberculosis.

This article has an online data supplement, which is accessible from this issue’s table of content online at www.atsjournals.org

## INTRODUCTION

Tuberculosis (TB) continues to be a major cause of morbidity and mortality worldwide. Standardized dosing of anti-tubercular drugs is effective in tuberculosis treatment and prevention, but may result in variable plasma drug levels and risk serious drug-related toxicities^1,2^. Studies have shown that a substantial proportion of patients treated for active TB experience at least one type of ADR (35%-68%), treatment failure (3%) or relapse (6-10%) within two years^3,4,5,6^. Liver enzyme elevations and drug-induced liver injury (DILI) are the most common adverse effects, affecting up to 30% of patients undergoing standard therapy^7,8,9,10^. Interindividual drug pharmacokinetic (PK) variation also affects treatment response. In a clinical cohort in South Africa, individuals who had any plasma drug concentration below target levels had 14-fold increased risk of microbiological failure, death, or relapse^11,12,13^.

A growing body of literature has identified mutations in genes encoding anti-TB drug-metabolizing enzymes that explain substantial PK variation and predict treatment outcomes and risk of adverse events.^14 15^. Mutations in N-acetyltransferase-2 (*NAT2*) and cytochrome P450 2E1 (*CYP2E1*) genes are known to affect metabolism and clearance of isoniazid^16^. Polymorphisms in the *NAT2* gene explain up to 88% interindividual pharmacokinetic variability of INH^17,18^. Based on mutations in the *NAT2* gene, individuals can be classified into three phenotypes—rapid, intermediate, and slow acetylators. Rapid acetylators typically have the lowest, while slow acetylators have highest plasma INH concentrations^19^. *CYP2E1* gene brings about conversion of acetyl hydrazine to reactive metabolites, which may result in hepatotoxicity^20^. Patients with *CYP2E1 Rsa*I polymorphism are significantly less likely to experience hepatotoxicity than those with the wild-type (*1A/*1A) genotype^21,22^. Associations between RIF clearance and mutations in drug transporter gene *SLCO1B1* and arylacetamide deacetylase *(AADAC)* have also been reported^23,24^. A study in South Africa found that patients with mutation in the *SLCO1B1* gene decreased RIF AUC.^25^ Mutations in cytochrome P450 gene *CYP3A5* are associated with faster LZD clearance, putting patients at higher risk of underexposure when treated at standard dose ^26,27^. In another study on South Africans treated for drug-resistant TB, CYP3A5*3 haplotype was associated with slower BDQ clearance^28^.

Modification of anti-TB drug doses based on pharmacogenomic data can improve PK target attainment, reduce toxicity risk, and improve treatment outcomes. Observational studies have shown that INH dose modifications enabled rapid and slow acetylators to achieve INH AUC targets comparable to those of intermediate acetylators^29^. A randomized trial of PGx-guided INH dosing among patients with active TB found that DILI was eliminated in slow acetylators (0% versus 78% in the standard dosing arm) and early treatment failure was reduced in rapid acetylators (15 vs 40% in standard dosing arm).^30^

Despite this evidence, pharmacogenomic-guided dosing is not widely used in treatment of TB. A major barrier to its implementation is the lack of scalable assays that can be performed quickly in facilities where TB is treated. At present, pharmacogenomic testing for *NAT2* and other relevant genes is typically only available in select reference laboratories and is often performed using expensive equipment that is not widely available in clinical laboratories, particularly in low- and middle-income countries where the majority of TB cases occur. In this study, we developed and validated a multiplex targeted sequencing-based panel to detect pharmacogenomic markers for INH, RIF, LZD and BDQ for use on Nanopore MinION sequencers, which are low-cost instruments that are increasingly accessible worldwide. We further validated our panel in a cohort of patients with active TB undergoing treatment, demonstrating utility to identify pharmacogenomic determinants of drug metabolism.

## METHODS

### Selection of pharmacogenomic markers

We searched published literature for pharmacogenetic markers of metabolism for drugs that are recommended by the World Health Organization for treatment of TB, including multidrug-resistant or RIF-resistant TB (MDR/RR-TB). We selected 15 well characterized single nucleotide polymorphisms (SNP) for which high quality studies had demonstrated associations with pharmacokinetic parameters, adverse events, or treatment outcomes^18–28^. The 15 SNPs, which had pharmacogenomic associations with INH, RIF, LZD or BDQ, occurred in five genes: N-acetyltransferase 2 (*NAT2),* cytochrome P450 family 2 subfamily E member 1 (*CYP2E1*), solute carrier organic anion transporter family member 1B1 (*SLCO1B1*), arylacetamide deacetylase *(AADAC)* and cytochrome P450 family 3 subfamily A member 5 (*CYP3A5*)^31^. These variations included 10 single nucleotide polymorphisms (SNP) located in exons, three in introns and two upstream of an exon **(Figure 1A)**.

**Figure 1.**
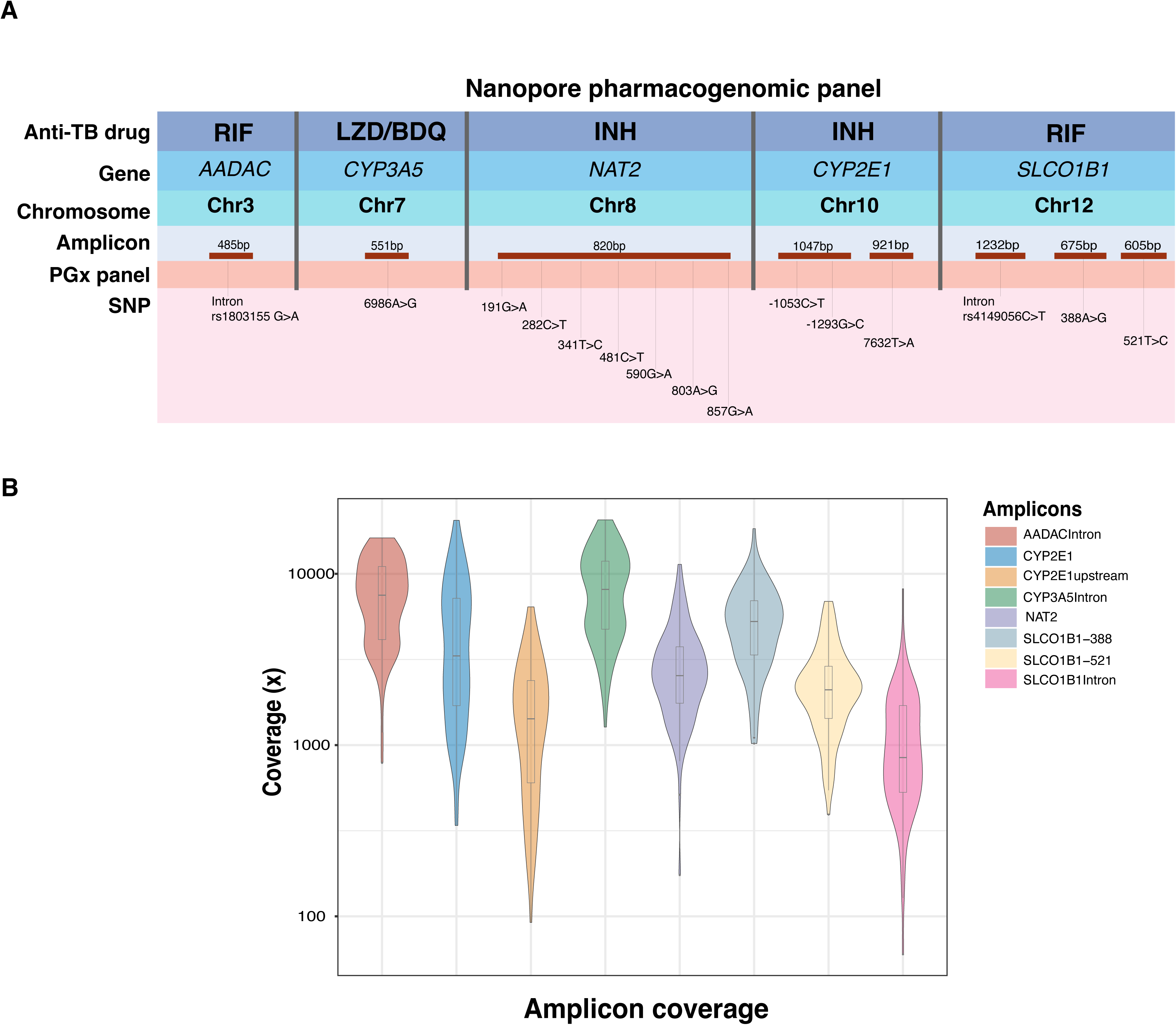
(A) Nanopore PGx panel. The top row contains the anti-TB drugs for which pharmacogenomic associations were identified from published studies. Genes and their location on chromosome are listed in rows two and three. Based on the position of targeted SNPs, targets were divided into eight amplicons corresponding to five genes (red). The amplicons are not scaled to their product length. The last row contains information on the positions in pharmacogenes which were included in the PGx panel. **(B) Amplicon coverage in PGx panel**: Sequencing coverage (log_10_ scale) per amplicon in 100 samples from the PK cohort, sequenced on a MinION Mk1C sequencer in a single-tube reaction.

### Primer design

To perform targeted sequencing, we used a multiplex strategy that relied on anchored primers We developed an 8-plex panel amplifying regions in five genes. Primers were designed to amplify products between 485bp–1232bp using Beacon Designer. (**Supplemental methods, Table E1**).

### DNA samples for panel development and validation

For the Nanopore pharmacogenomic panel development and validation, 48 purified DNA samples from the 1000 Genomes Project for which Illumina whole genome sequencing data was available were procured from the Coriell Institute for Medical Research, USA^31^. Additionally, we sequenced oral swabs from healthy volunteers (n=20) using custom PGx panel (**supplemental methods**).

### Single-tube multiplex PCR

A single-tube 8-plex PCR reaction was performed for each sample using the LongAmp® Taq DNA Polymerase (NEB). The final reaction was carried out in a 50μl volume containing 1x LongAmp Taq Reaction Buffer (NEB), 2.5M Betaine (Sigma), 5U LongAmp Taq DNA polymerase. Primer efficiency was first evaluated on a single-plex reaction with subsequent addition of primer sets to evaluate the non-specific binding and inhibition of each primer set by the others. The final PCR conditions were optimized to 2 min of DNA denaturation at 94 °C followed by 30 cycles of amplification as follows: 30 sec at 94 °C for denaturing, 60 sec at 60 °C for primer annealing, 1.5min at 65 °C for extension, followed by 10 min at 65 °C for a final extension. The PCR product was purified with PureLink™ PCR Purification Kit (Thermo Fisher Scientific). The purified PCR product was eluted with 50μl nuclease-free water and quantified with Qubit to evaluate reaction yield.

### MinION library preparation and sequencing

For panel development and validation, we sequenced a total of 60 Coriell DNA samples (one Coriell DNA sample at 12 dilutions and additional 48 Coriell DNA samples with different genotypes) and 20 oral swabs samples from healthy individuals. The library was prepared using the SQK-LSK110 Ligation Sequencing Kit (Oxford Nanopore Technologies). Samples were barcoded using a Nanopore PCR barcoding expansion (EXP-PBC096 PCR Barcoding Expansion)followed by DNA repair and end-prep using NEBNext FFPE DNA Repair Mix and NEBNext Ultra II End repair / dA-tailing Module reagents in accordance with manufacturer’s instructions. Adaptor ligation was performed using Adapter Mix F (AMX-F) and Quick T4 Ligase. The samples were sequenced on a MinION Mk1C sequencer (**Supplemental methods, Table E2**).

### Sequencing data analysis

Demultiplexing and real-time basecalling was performed on an in-built software MinKNOW (release 22.08.4) using an onboard basecalling software Guppy (Version 3). The run was set on a high accuracy base calling (cutoff >9). Mapping was done aligning reads to a multi-fasta file containing the concatenated sequences of the genome regions included in the panel. Reference fasta genome was uploaded on Epi2ME (version 4.1.3.) cloud and fastq files with passed reads well aligned to the custom genome to generate bam files. To call variants first, the reads were mapped to the reference sequences of target genes included in the gene panel using Minimap2 (v2.26) with default parameters. Aligned reads with a mapping quality score under 60 (MAPQ60) were discarded. Variant calling was performed with Calir3 using default ONT settings. Variants identified were phased using WhatsHap (v1.7).

### Samples for clinical validation Study cohort and ethical approval

Patients with GeneXpert MTB/RIF-confirmed RIF-susceptible pulmonary TB were recruited at the Ubuntu HIV/TB Clinic, Site B, Khayelitsha, South Africa (University of Cape Town Faculty of Health Sciences Human Research Ethics Committee approval 568/2012) as a part of a larger study. Whole blood samples (n=100) from a subcohort of this study who were invited to participate in a nested pharmacokinetic study between July 2013 and April 2014 were used for pharmacogenomic validation^32^. All patients provided written consent prior to participation. Detailed sociodemographic data, past TB treatment history, and comorbidity data were collected. Weight band-based dosing was used in line with WHO guidelines (**Table 1**).

**Table 1.** Clinical characteristics of tuberculosis pharmacokinetic cohort.

|  | PK cohort<br>(N= 100) |
| --- | --- |
| **Clinical characteristic** |  |
| Female sex | 43 |
| Age in years, median (IQR) | 33 (29–40) |
| Smear grade at baseline |  |
| 3+ | 24 |
| 2+ | 22 |
| 1+ | 20 |
| Scanty/Negative | 34 |
| Baseline time to culture positivity (days), median (IQR) | 10 (7–14) |
| Extensive radiological disease at baseline | 71 |
| Cavities at baseline | 52 |
| Smoking history |  |
| Current | 24 |
| Previous | 27 |
| Never | 49 |
| Alcohol consumption | 37 |
| Retreatment | 39 |
| Type 2 diabetes mellitus | 4 |
| BMI at PK study (kg/m<sup>2</sup>), median (IQR) | 21.5 (20–23) |
| Albumin concentration at PK study (g/liter), median (IQR) | 38 (34–40) |
| Dose (mg/kg), median (range) |  |
| Rifampin | 10 (7–11.5) |
| Isoniazid | 5 (3.5–6) |
| Participants reporting side effects of TB treatment | 35 (35) |

Numerical values reflect number of participants with the characteristic unless otherwise specified.

### Pharmacokinetic data

A description of the sampling, therapeutic drug monitoring and pharmacokinetic analysis for this cohort has previously published^32^. Briefly, pharmacokinetic sampling was carried out for RIF and INH 7 to 8 weeks after initiation of anti-TB therapy. Blood samples were obtained immediately before (pre-dose) and 1, 2, 3, 4, 6, and 8 hours after drug ingestion. They were immediately placed on ice, and plasma was separated by centrifugation within 30 minutes before storage at −80°C until analysis. The storage tubes containing the plasma samples were transferred on dry ice to the Clinical Pharmacology Laboratory at the University of Cape Town, where drug concentrations were determined using validated liquid chromatography tandem mass spectrometry (LC-MS/MS) methods^35^.

Whole blood samples from 100 TB positive patients for which pharmacokinetic data was available were collected in citrate tubes and stored in −80°C until used. DNA was extracted from 100ul whole blood samples using DNeasy Blood & Tissue Kit (Qiagen) and eluted in 50ul nuclease-free water. Approximately 50ng of purified DNA was used for targeted sequencing using custom Nanopore PGx panel as described above.

### Haplotype labelling

For INH pharmacogenomic analysis, phased *NAT2* haplotypes for PK cohort derived from Nanopore sequencing were labelled based on seven canonical SNPs following an international consensus nomenclature to interpret acetylator phenotype^33^. The *CYP2E* allele nomenclature was quoted based on the Human Cytochrome P450 Allele Nomenclature Committee tables^34^

### Statistical analyses

Previously developed population pharmacokinetic models were used to test the effect of polymorphisms or acetylator type on clearance and bioavailability within the models using Monolix (Version 2023R1; Lixoft SAS)^35^. Stepwise covariate selection was performed using a drop in objective function value of >3.84 as a cutoff for inclusion (corresponding with a p<0.05) and an increase of >6.64 as a cutoff for backward elimination (p<0.01). Pre- and postprocessing of data was done in R (Version 4.3.1).

## RESULTS

### PGx panel performance and coverage per amplicon

To evaluate the panel’s sensitivity and accuracy, we first performed targeted Nanopore sequencing on six samples in duplicates that were obtained by diluting a Coriell DNA sample with known genotype. At all dilutions, we obtained median coverage above the minimum cutoff (>50x). Coverage from 500ng to 50ng was 850x, 847x, 972x, 1005x, 1066x and 1076x respectively. The median quality score of diluted samples was 12.8 (std dev=0.1), and the median yield for passed reads was 15Mb (std dev=2.4). (**Figure E1)**. We then validated the PGx panel on 48 purified DNA samples from Coriell Institute. Majority of the Coriell samples selected for panel validation were from sub-Saharan Africa (54.1%) and the Americas (25%) and consisted of 41.3% males. We achieved complete coverage of the targeted regions by aligning eight PCR amplicons in the PGx panel. All amplicons were sequenced with coverage depth above the minimum cutoff. The median sequencing depth across eight amplicons in Coriell samples was ∼ 2,281x, with 99.7% amplicons above 100x and 90.2% above 500x. Among the eight amplicons, CYP3A5Intron had the highest coverage (median= 6934x; IQR= 2,522-9,534.7), while SLCO1B1Intron had the lowest coverage (median= 779; IQR= 429.5-1,221.5). We observed 100% concordance between variants identified in Nanopore PGx panel and the reference Illumina whole genome sequencing.

We also sequenced 20 DNA samples extracted from oral swabs collected from healthy volunteers to demonstrate the use of oral swab as an alternate non-invasive sampling method for pharmacogenomic testing. We obtained high quality sequencing data for all oral swab samples with the majority of amplicons above 100x coverage (96.8%).

### Clinical validation

For clinical validation, we performed targeted Nanopore sequencing on DNA extracted from whole blood samples from active TB patients enrolled in the INH and RIF PK cohort (**Table 1**). A majority of the participants were of Xhosa ethnicity 98/100 (98%) and 65% of the participants was living with HIV. The median age was 33 years (range= 29–40), and 43% were women. Median quality score of the samples was 13.8 (IQR=13.5-14.0), We obtained full coverage of the targeted regions for every sample, with a coverage depth that exceeded the minimum cutoff (>50x) for all amplicons. The median sequencing depth across eight amplicons in the PK cohort was ∼ 2,963x (IQR 1,512-6,156), with 99.8% amplicons above 100x and 93.6% above 500x. (**Figure 1B**).

### INH and RIF Pharmacogenomic associations

We obtained read depth above 100x for variant alleles at all positions. A total 253 homozygous mutant and 353 heterozygous alleles were detected in 100 samples at 15 genomic positions. The frequency of homozygous wildtype, homozygous alternate and heterozygous variant alleles is shown in **Figure 2A** and **Table 2**.

**Figure 2.**
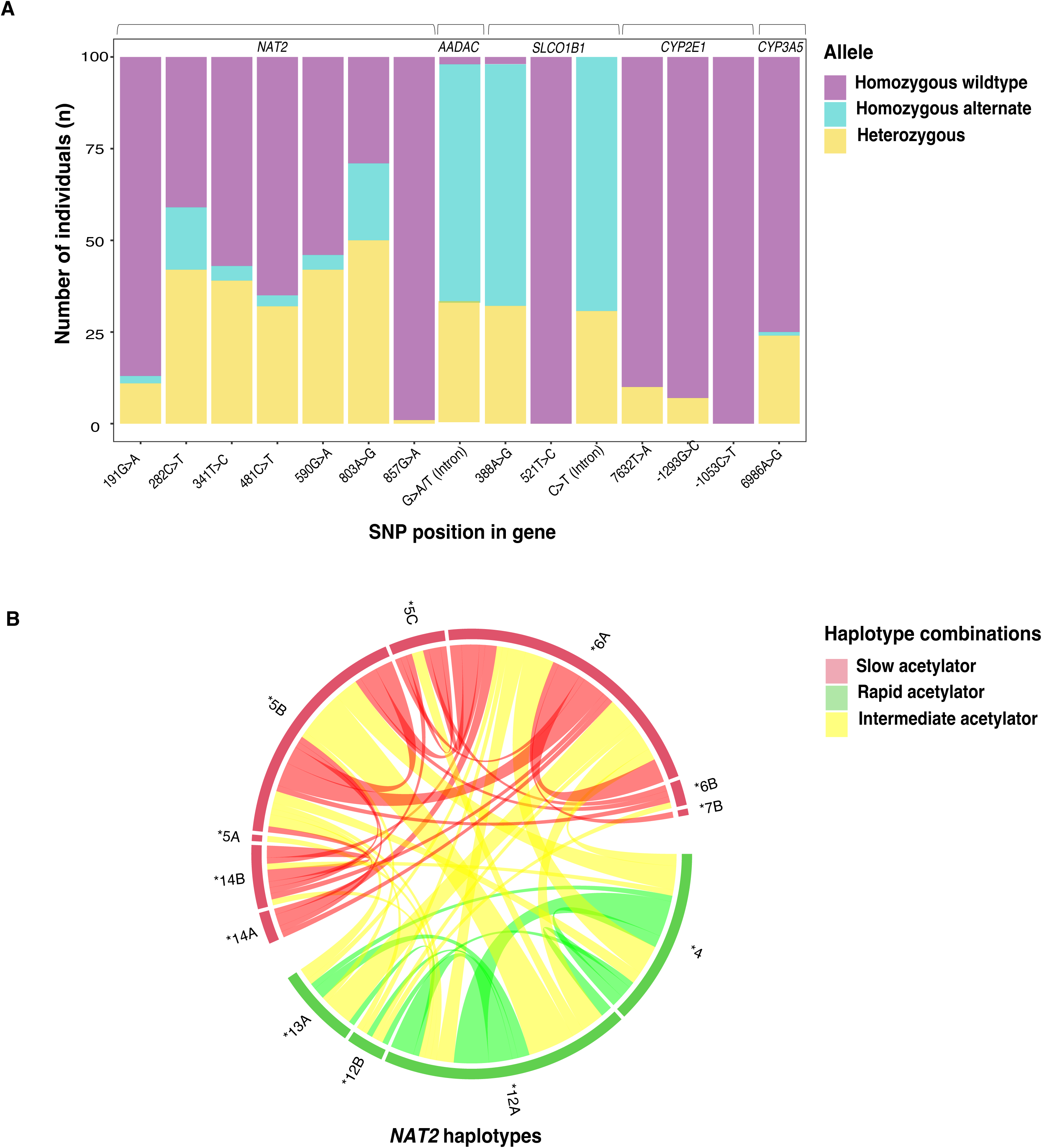
**(A)** Distribution of homozygous wildtype (purple), homozygous alternate (blue) and heterozygous alleles (yellow) at 15 polymorphic sites in active TB patients (n=100) from PK cohort sequenced on MinION sequencer. **(B)** *NAT2* haplotypes in red are slow acetylator types, those in green are rapid acetylator haplotypes. Connections in red indicate two slow acetylator haplotypes, those in green indicate two rapid haplotypes, and those in yellow indicate one rapid and one slow haplotype (intermediate acetylation)

**Table 2.** Variant calling summary of fifteen PGx panel markers for 100 clinical cohort samples analyzed on a Nanopore MinION sequencer.

| Pharmacogene | SNP identifier | SNP | WT<sup>#</sup> (%) | MUT* (%) | HET<sup>\$</sup> (%) | Genotype quality, median | Read depth, median |
| --- | --- | --- | --- | --- | --- | --- | --- |
| *NAT2* | rs1801279 | 191G>A | 87 | 2 | 11 | 20.0 | 4801.0 |
| *NAT2* | rs1041983 | 282C>T | 41 | 17 | 42 | 22.0 | 5180.0 |
| *NAT2* | rs1801280 | 341T>C | 57 | 4 | 39 | 23.0 | 4775.0 |
| *NAT2* | rs1799929 | 481C>T | 65 | 3 | 32 | 21 | 4976 |
| *NAT2* | rs1799930 | 590G>A | 54 | 4 | 42 | 21.0 | 5339.0 |
| *NAT2* | rs1208 | 803A>G | 29 | 21 | 50 | 20.0 | 4715.0 |
| *NAT2* | rs1799931 | 857G>A | 99 | 0 | 1 | 23.0 | 6457.0 |
| *CYP2E1* | rs3813867 | -1293G>C | 93 | 0 | 7 | 21.0 | 5377.0 |
| *CYP2E1* | rs2031920 | -1053C>T | 100 | 0 | 0 | NA | NA |
| *CYP2E1* | rs6413432 | 7632T>A | 90 | 0 | 10 | 20.0 | 8377.0 |
| *SLCO1B1* | rs4149032 | C>T | 0 | 70 | 30 | 24.0 | 1721.0 |
| *SLCO1B1* | rs4149056 | 521T>C | 100 | 0 | 0 | NA | NA |
| *SLCO1B1* | rs2306283 | 388A>G | 2 | 66 | 32 | 24.0 | 4979.0 |
| *AADAC* | rs1803155 | G>A/T | 2 | 65 | 33 | 23.0 | 8047.0 |
| *CYP3A5* | rs776746 | 6986A>G | 75 | 1 | 24 | 21.0 | 8053.0 |

Based on the international consensus nomenclature, participants were classified as slow (33/100; 33%), intermediate (47/100; 47%) and rapid (20/100; 20%) acetylators. Demographic and clinical characteristics did not differ by acetylator type (**Table 3**). The *NAT2* haplotype distribution for 100 PK samples is provided in **Figure 2B**. INH clearance rates were lowest in slow acetylators (median,11.2 L/h; IQR, 9.3-13.4), moderate in intermediate acetylators (27.2 L/h; 22.0-31.7), and highest in fast acetylators (45.1 L/h; 34.1-51.1) (**Table 3**; **Figure 3A**). The area under the plasma drug concentration-time curve (AUC) for 0-24h was lowest for rapid acetylators (median 5.80 mg*h/L; IQR, 4.38-9.48), moderate for intermediate acetylators (10.7 mg*h/L; 7.94-14.6) and highest in slow acetylators (23.2 mg*h/L; 18.3-30.9) (**Table 3**; **Figure 3B**). *NAT2* acetylator status had a significant effect on INH clearance (dOFV=105.5, p<0.0001) (**Figure E2)**. In addition, the effect of HIV, that was previously in the model, was now no longer significant after backward elimination. Individuals who were slow acetylators were more likely to report side effects than intermediate or fast acetylators (52% vs 27%, p=0.027).

**Figure 3.**
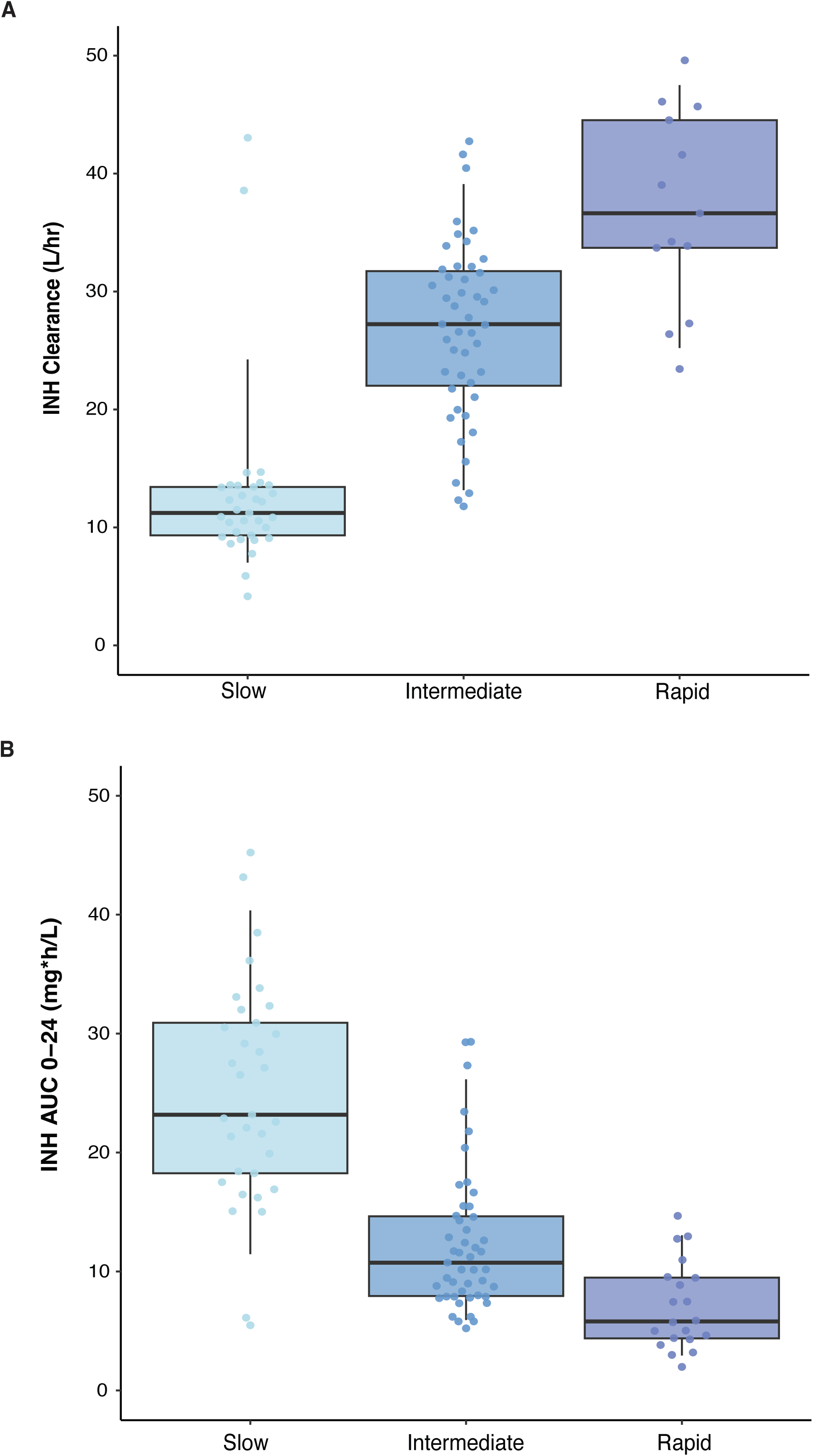
Predicted phenotype and INH clearance. (**A)** Predicted *NAT2* phenotype and INH clearance in active TB patients (n=100) rapid acetylators (purple) retained plasma drug levels for a shorter period than intermediate (blue) and slow (light blue) predicted phenotypes (p-value= <0.0001). **(B) The area under the plasma drug concentration-time curve (AUC) for 0-24hrs** was lowest for rapid acetylators (5.80 (4.38-9.48) mg*h/L), moderate for intermediate acetylators (10.7 (7.94-14.6) mg*h/L) and highest in slow acetylators (23.2 (18.3-30.9) mg*h/L)

**Table 3.** Demographic and clinical characteristics, and INH pharmacokinetic parameter estimates, by *NAT2* acetylator status.

|  | **Slow**<br>**(n=33)** | **Intermediate**<br>**(n=47)** | **Rapid**<br>**(n=20)** | **p-value** |
| --- | --- | --- | --- | --- |
| **Age (years)** | 33.4 (28.9-39.0) | 32.4 (30.3-40.6) | 29.5 (27.1-41.6) | 0.44 |
| **Sex, female, n (%)** | 16 (48) | 19 (40) | 8 (40) | 0.739 |
| **Albumin** | 39 (36.0-42.0) | 39.0 (34.0-43.0) | 39.5 (36.3-76.3) | 0.785 |
| **Days on TB treatment, median (IQR)** | 56.0 (53.0-57.0) | 56.0 (52.0-60.5) | 54.5 (48.8-58.0) | 0.455 |
| **HIV infected, n (%)** | 8 (24) | 19 (40) | 8 (40) | 0.286 |
| **BMI (kg/m<sup>2</sup>), median (IQR)** | 22 (19.3-23.0) | 22.0 (19.3-23.0) | 21.5 (19.8-23.0) | 0.532 |
| **Isoniazid PK, median (IQR)** |  |  |  |  |
| Clearance (L/hr) | 11.1 (9.2, 12.9) | 26.2 (20.5, 32.1) | 42.0 (33.5, 52.8) | <0.0001 |
| AUC (mg*hr/L) | 23.1 (18.2, 30.3) | 10.7 (7.9, 14.6) | 5.9 (4.4, 9.5) | <0.0001 |
| Cmax (mg/L) | 4.5 (3.4, 5.5) | 3.3 (2.6, 4.5) | 2.4 (1.7, 3.5) | <0.0001 |

We also evaluated whether polymorphisms in *CYP2E1*, part of the downstream INH metabolism pathway, could explain pharmacokinetic variability or were associated with adverse events. Eighty-seven participants were *1A/1A (wildtype) genotype, 7 were *5B/*5A and 6 were *1A/*6. After inclusion of acetylator status, 7632T>A (rs6413432) had a significant effect on INH bioavailability. Participants who were heterozygotes (n=10) had a 23% (2.2-50) higher bioavailability than wild type patients (p = 0.0008). We did not observe any significant associations (p=0.28) between *CYP2E1* haplotypes and reported side effects.

For RIF pharmacogenomic analysis, we analyzed three SNP sites in *SLCO1B1* (rs4149032 C>T, 388A>G and 521T>C) and one in *AADAC* (rs1803155). At the rs4149032 position, 70/100 patients were homozygous mutant and 30/100 were heterozygous alleles. We identified 2/100 wildtype, 66/100 homozygous alternate 32/100 heterozygous alleles at 388A>G position. All samples were detected as wild type at 521 T>C position in *SLCO1B1* gene. We did not observe any significant associations between *SLCO1B1* mutations and RIF bioavailability. RIF clearance was 16.5% (1.30-29.3) lower in individuals who were homozygous alternate for *AADAC* rs1803155 G>A substitutions (p=0.0015; **Figure 4, Figure E3).**

**Figure 4.**
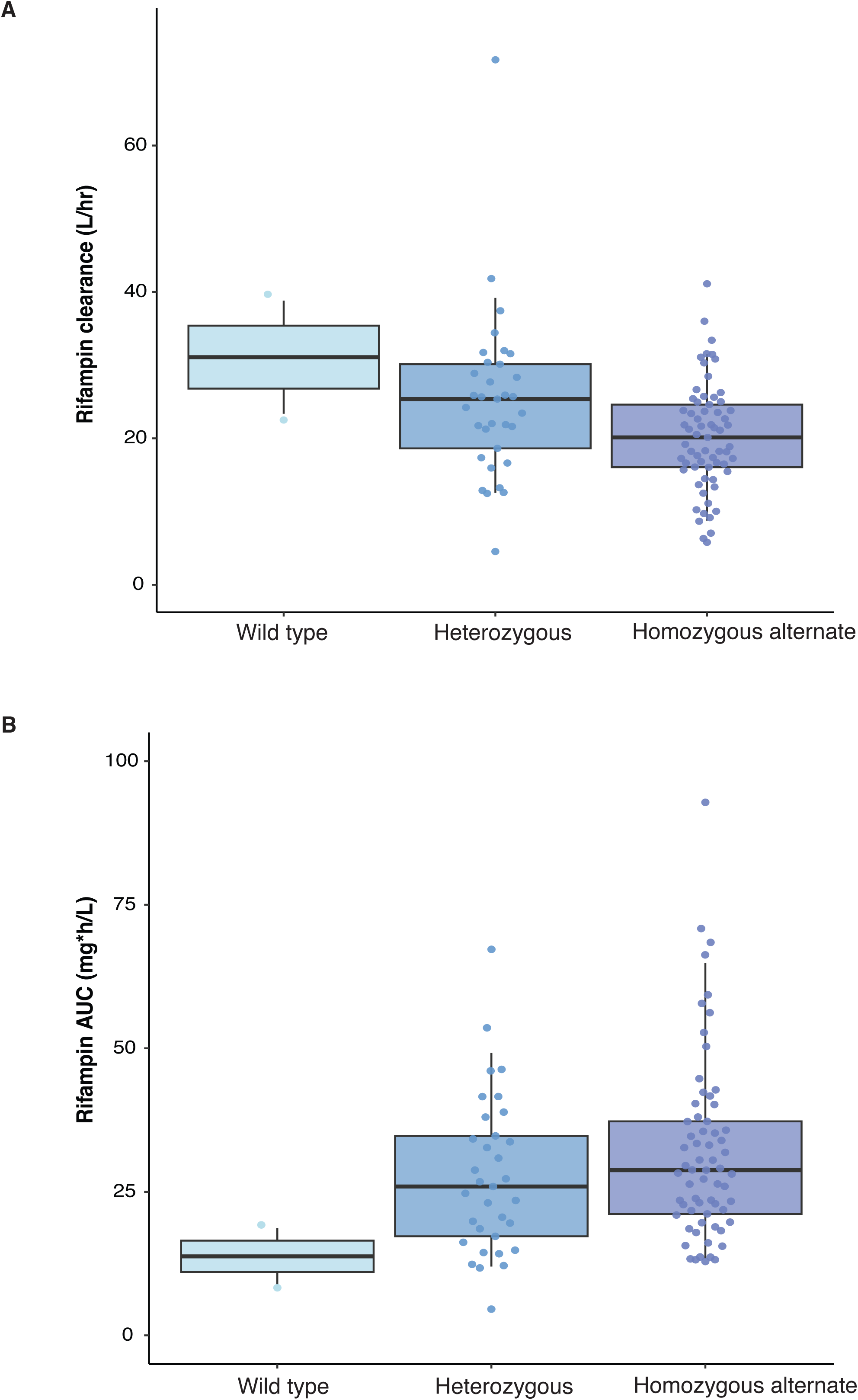
(A) *AADAC* rs1803155 G>T intron mutation and effect on rifampicin clearance: RIF clearance was 16.5% (1.30-29.3) lower in individuals who were homozygous alternate (purple) for *AADAC* rs1803155 G>A substitutions (p=0.0015) than heterozygous (blue) and wild type (light blue) **(B) The area under the plasma drug concentration-time curve (AUC) for 0-24hrs in RIF:** Homozygous alternate (purple) for *AADAC* rs1803155 G>A substitutions heterozygous (blue) and wild type (light blue)

## DISCUSSION

While TB is treatable and preventable, a substantial proportion of patients experience drug associated toxicities, treatment failure and relapse under standardized dosing. For preventive therapy, adverse drug events, which are associated with drug metabolism, are a strong predictor of non-completion^36^. Pharmacogenomic guided dosing has the potential to reduce the risk of these poor outcomes, with observational studies and a randomized trial demonstrating strong premise for feasibility and effectiveness^50^. However, a major obstacle to using PGx-guided dosing is the lack of access to pharmacogenomic assays in clinical settings where TB is common. To address this gap, we developed a single-tube targeted sequencing panel on Oxford Nanopore MinION platform to detect mutations associated with the metabolism of INH, RIF, LZD and BDQ for which pharmacogenomic associations were previously reported. We achieved high coverage and read depth for all targets in the panel and found that variant identification was 100% concordant in well characterized reference genomes. As proof of principle, we performed the assays on samples from an active TB clinical cohort in Cape Town, confirming that *NAT2* acetylator types strongly predicted INH clearance in this population.

Currently available methods used for detection of mutations in pharmacogenes largely rely on qPCR, restriction fragment length polymorphisms (RFLP), SNP array platforms, single gene Sanger sequencing or larger scale (exome or whole genome) sequencing^37^. While qPCR methods are rapid and easier to perform, they target only limited number of mutations and provide unphased data and *in silico* haplotype predictions. One consequence of this is that polymorphisms that important in some populations are sometimes neglected. For example, the G191A (R64Q) SNP is common to the NAT2*14 allele cluster, which is frequent in African and African American individuals but is rarely observed in other populations^38^, leading it to be left out of the popular *NAT2* phasing tool *nat2pred* ^39^. One study found no correlation between *NAT2* genotype and INH metabolism in individuals of Zulu descent, South Africa; however, the study excluded G191A SNP, leading to a population-specific prediction bias ^40^. SNP array platforms and whole exome/genome sequencing provide data covering more genes and relevant SNP, but typically require expensive laboratory equipment and are not widely available in clinical facilities in resource-constrained settings.

We previously developed a qPCR-based pharmacogenomic assay on the GeneXpert platform to detect polymorphisms in *NAT2* gene to guide INH dosing^41^. This assay predicted INH metabolism with high accuracy based on 5 canonical SNPs; however, there are constraints to including further targets for a single-tube cartridge-based assay. As TB treatment requires at least three drugs and many drugs have several relevant pharmacogenes and multiple important SNP per gene, an optimal panel will require multiple targets. We identified 15 SNP in 5 genes for which there are compelling pharmacogenomic data for important anti-TB drugs. Our search identified many other SNPs for which data were sparse or conflicting; as further studies confirm or reject associations between these polymorphisms, our assay could be easily expanded to include other targets.

In the current study, which covered all 7 canonical SNPs in *NAT2,* we found that *NAT2* haplotypes were strongly predictive of isoniazid clearance and AUC; clearance was nearly 4 times higher, and AUC 4 times lower, in rapid compared with slow acetylators. Prior studies have demonstrated that increasing isoniazid dosing among rapid acetylators (to 7.5-10 mg/kg) and decreasing it among slow acetylators (to 2.5 mg/kg) can achieve PK targets and reduce adverse events^29^. Given the diversity of *NAT2* acetylator types in this population and globally, testing combined with isoniazid dose modification could confer substantial clinical benefits^42^. We found a modest effect of rs1803155 G>A substitution in the arylacetamide deacetylase gene, *AADAC*; homozygous individuals had 17.3% lower clearance than heterozygous and wild type alleles (p=0.0015). AADAC appears to be main enzyme in rifampin deacetylation^43^, and studies have found it affects rifamycin exposure. Chigutsa et al. previously reported that *SLCO1B1* rs4149032 polymorphism is highly prevalent in South Africans and is associated with reduced rifampin concentrations^23^. In our study, all participants were heterozygous or homozygous alternate for the rs4149032 polymorphism, so we lacked a reference group of wild type individuals for comparison. The high frequency of rs4149032 polymorphisms in this population, together with data showing that low rifampin AUC is predictive of poor outcomes, add to the growing evidence that higher doses of rifampin may be needed^14^.

We included pharmacogenetic targets associated with LZD and BDQ toxicity, though these are not first line TB drugs. LZD and BDQ are now both included in the primary WHO recommended regimen for treatment of MDR/RR-TB ^44^. Additionally, a recent trial demonstrated that an 8-week course including LZD and BDQ for RIF-susceptible TB was non-inferior to standard 6-month therapy, opening the door to potential ultra-short-course regimens^45^. Both drugs are associated with serious adverse events. Peripheral neuropathy and myelosuppression are common with prolonged courses of LZD and can be treatment limiting. QT prolongation, leading to serious arrythmias, can be seen with BDQ, particularly when used in combination with other QT prolonging drugs including moxifloxacin. Recent studies have found that polymorphisms in *CYP3A5* may influence LZD and BDQ clearance. The *CYP3A5 *1* haplotype was associated with a nearly six-fold risk of LZD underexposure compared with *3/*3, and the *3 haplotype was associated with slower clearance of BDQ, including 30% lower clearance for homozygous individuals (*3/*3)^26,27,28^. While we analytically validated our assay to correctly identify these polymorphisms, our clinical cohort did not include individuals receiving these drugs, and further studies are needed to confirm the importance of these variants in diverse populations.

Amplicon-based approaches coupled with MinION sequencing offers several advantages over conventional methods. MinION supports real-time base calling that allows users to stop or pause the run when the output is enough for the analysis. This is highly advantageous when used in clinical settings where quick results are needed to decide treatment, improve prognosis, and guide clinical management. A recent study employed a custom variant-prioritization approach with Nanopore sequencing to rapidly diagnose various disorders in critically ill patients^46^. In the present study, we performed 24 hour and 48-hour runs, resulting in coverage that was several-fold higher than the required cutoff value. Based on these findings, the run time of the PGx assay can be cut down to a few hours. Targeted Nanopore sequencing has been previously used to detect drug resistant strains in MTB from sputum^47^ and point-of-care diagnosis of viral and bacterial infections^48,59^. Another advantage of using the Nanopore sequencing approach is availability of a smaller and cheaper Flongle flowcells (∼$90), which produce up to 2.8 Gb of output. We found that we could sequence 50 samples on each Flongle run with sufficient coverage. Since targeted sequencing provides information on the entire gene sequence or the targeted amplicons, novel mutations can be identified in these targets which can provide valuable insights into the evolution of these genes in different populations. Our assay requires single tube PCR amplification and library preparation prior to sequencing on Nanopore; instruments to automate these processes are becoming available, which will be important for amplicon sequencing assays to be implemented in clinical laboratories.

The findings of this study are subject to several limitations. To increase the efficiency of the multiplex assay, we split the target genes into two or more amplicons covering a region of the gene instead of the full length. Due to this, we may have missed novel mutations in those regions. Although, we achieved high coverage for all amplicons, there was moderate variability in coverage across the amplicons. Furthermore, while we developed the custom panel for four anti-TB drugs, PK data was available only for INH and RIF to predict pharmacogenomic associations, and there was a lack of genetic diversity at some sites that had been previously identified as important for metabolism of these drugs. Further studies are needed in diverse populations and to assess the impact of *CYP3A5* mutations in LZD and BDQ metabolism. Liver enzyme data were not available to assess the effect of *CYP2E1* mutations on drug-induced liver injury, which has been previously reported in several studies^22^.

Amid growing evidence that we can identify individuals at greatest risk of anti-TB drug toxicities and poor treatment outcomes by screening for common genetic variants, there is a need for assays that can be performed near to the point of clinical care in settings where TB is common. We developed and validated a Nanopore amplicon sequencing panel to detect pharmacogenomic markers for key first- and second-line anti-TB drugs. This panel can be further expanded as additional pharmacogenetic markers of TB medications are identified and validated. This assay can be performed on MinION devices, which cost around $1,000 and are increasingly available in public health laboratories in low- and middle-income countries. The movement to optimize TB treatment for each patient will require tools such as this that are scalable for use in settings where TB burden is greatest.

## Data availability

Data supporting the findings of this manuscript are available in the Supplementary Information files or from the corresponding author upon request.

## Author contributions

R.V. and J.R.A. conceived of the study. R.V., J.R.A. and R.E.W wrote the first draft of the manuscript. R.V., J.R.A, N.R. and R.J.W. designed the experiments. N.R., and R.J.W. recruited patients and collected patient’s data. R.E.W, N.R. and P.D. performed PK analysis. R.V., N.Y., T.S., E.K. and K.dS performed the experiments. R.V., K.dS., J.R.A and R.E.W analyzed data. All authors contributed to the final version of the manuscript.

## At a Glance Commentary

#### Scientific Knowledge on the Subject

Standardized dosing of anti-tubercular (TB) drugs results in variable plasma drug levels, which is associated with adverse drug reactions, poor treatment outcomes, and a risk of relapse. Mutations in genes affecting drug metabolism may explain this pharmacokinetic variability; however, pharmacogenomic (PGx) assays that predict metabolism of anti-TB drugs have been lacking.

#### What This Study Adds to the Field

We developed a custom single-tube Nanopore sequencing panel to detect mutations for predicting the metabolism of isoniazid, rifampicin, linezolid, and bedaquiline. Such assays are not currently available in clinical settings to guide drug dosing. We validated our panel on Coriell DNA samples (n=48) and achieved 100% concordance with Illumina whole genome sequencing data. Next, we validated the predicted metabolism of isoniazid and rifampicin based on genotypes derived from the PGx panel in patients with active TB (n=100) undergoing treatment and found strong correlation with INH metabolism. Targeted sequencing on an affordable and portable device can facilitate the identification of polymorphisms that impact TB drug metabolism, allowing for personalized dosing in TB treatment or prevention.

---

**Figure E1.**
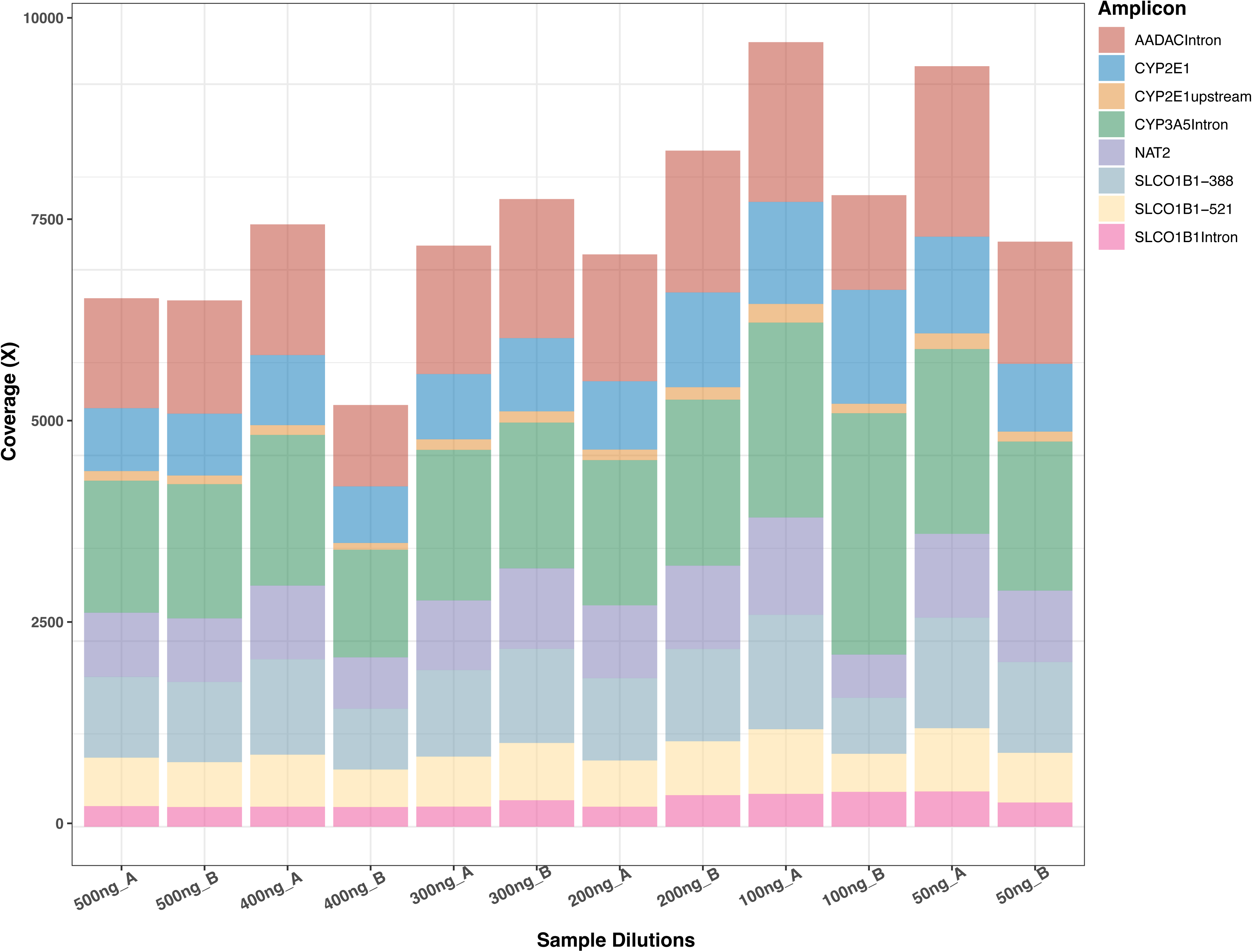
Coverage per amplicon at different dilutions: Amplicon sequencing coverage distribution in diluted samples. 1000 Genomes Project sample DNA sequenced at six dilutions in replicates from 500ng to 50ng.

**Figure E2.**
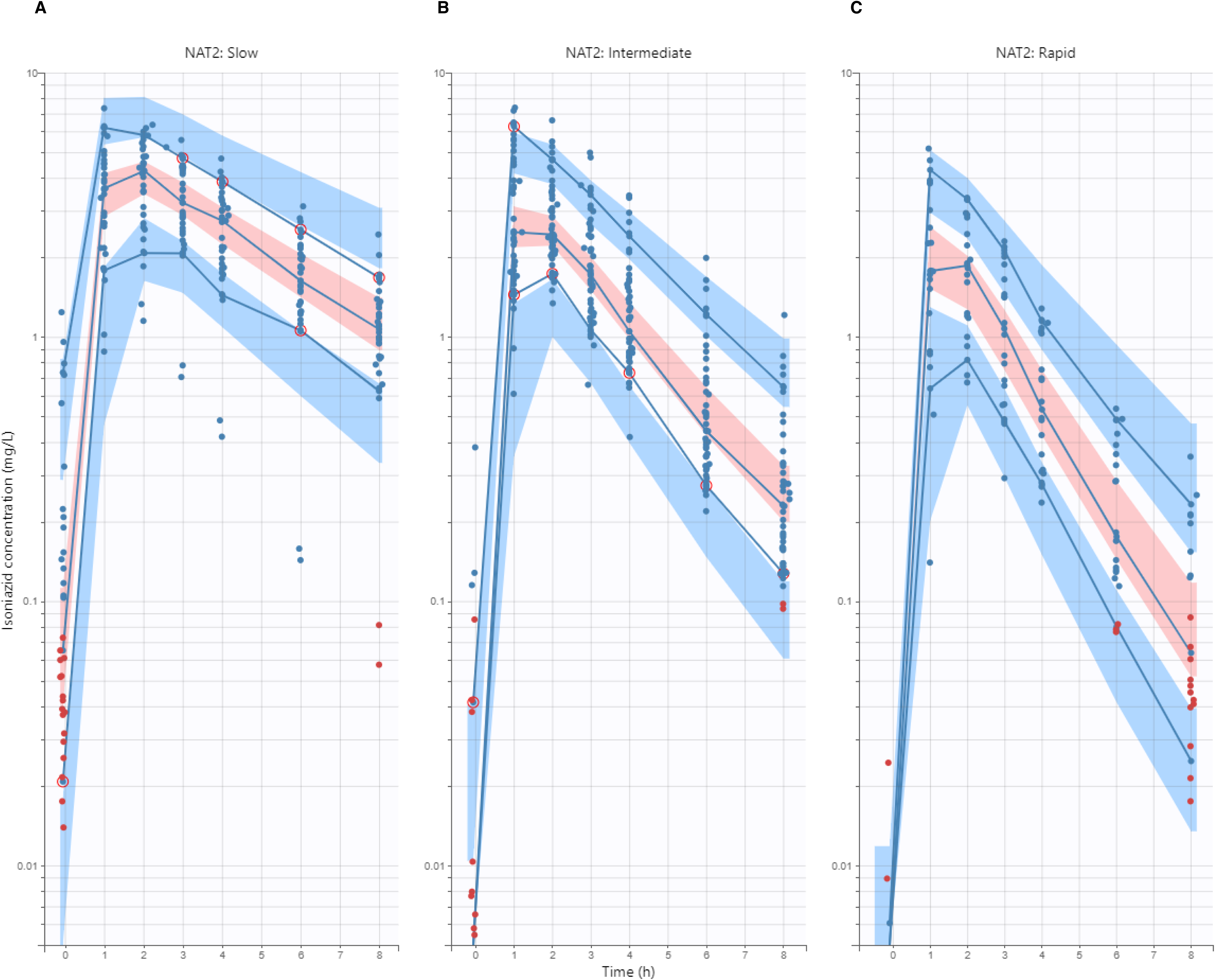
Visual predictive check (VPC) for isoniazid concentration (mg/L) versus time (hours), stratified by predicted NAT2 acetylator status **(A)** Slow acetylators **(B)** Intermediate acetylators **(C)** Rapid acetylators.

**Figure E3.**
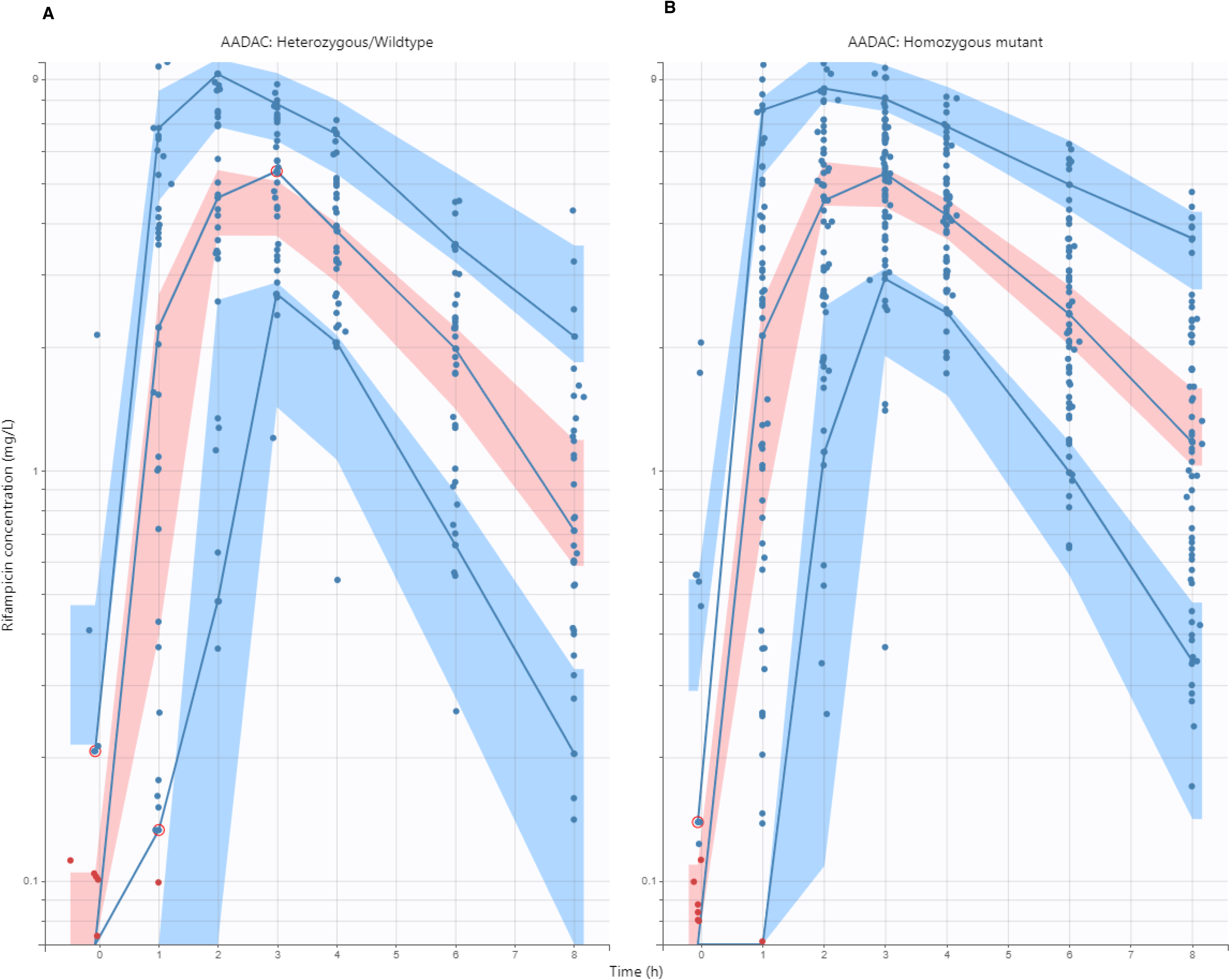
Visual predictive check (VPC) for rifampicin concentration (mg/L) versus time (hours), stratified by mutation at rs1803155 position in *AADAC* gene **(A)** Heterozygous for rs1803155 G>A mutation in *AADAC* gene. **(B)** Homozygous alternate for rs1803155 G>A mutation in *AADAC* gene.

