## Supplementary material for "A Nanopore sequencing-based pharmacogenomic panel to personalize tuberculosis drug dosing": Online supplement

**METHODS**

**Primer design**

To perform targeted sequencing, we used a multiplex strategy that relied on anchored primers. This involves amplifying the regions of interest using specific primers flanked by Nanopore compatible anchor sequences (5’-Anchor- TTTCTGTTGGTGCTGATATTGC and 3’-Anchor- ACTTGCCTGTCGCTCTATCTTC). The anchor sequences enable a second round of PCR with Oxford Nanopore's barcoded outer primers for rapid adapter attachment. The second PCR uses fewer cycles and creates barcoded amplicons with modified 5' ends for simplified post-PCR adapter attachment. We developed an 8-plex panel amplifying regions in five genes. Primers were designed to amplify products between 485bp–1232bp using Beacon Designer (Premier Biosoft International, version 8). Primer specificity was verified with Primer-blast (NCBI). Primer concentrations and sequence details are listed in **Supplementary table 1**.

**DNA samples for panel development and validation**

For the Nanopore pharmacogenomic panel development and validation, 48 purified DNA samples from the 1000 Genomes Project for which Illumina whole genome sequencing data was available were procured from the Coriell Institute for Medical Research, USA. Approximately 100ng of purified DNA from each sample was directly used for the 8-plex PCR. To assess the analytical sensitivity and variant calling accuracy of custom PGx panel, one Coriell DNA sample with known genotype was sequenced at six dilutions (50ng, 100ng, 200ng, 300ng, 400ng, 500ng) in replicate.

**DNA extraction from oral swabs**

Oral swabs were collected from 20 healthy volunteers at the Stanford School of Medicine, CA, USA. Each individual was given a sterile hyraflock swab (Longhorn Diagnostics, Maryland, US) and asked to collect buccal cheek swabs. The participants were instructed to rinse their mouth with water before swab collection to avoid any food particle contamination. The swabs were collected in 200ul of nucleic acid preservative and transport media (Primestore MTM, Longhorn Diagnostics, Maryland, US) and stored at -20°C until extraction. DNA was extracted from oral swabs using QIAamp DNA Mini Kit (Cat # 51304) per the manufacturer’s instructions. The samples were eluted in 50μl nuclease-free water and stored in -20°C until use.

**MinION library preparation and sequencing**

For panel development and validation, we sequenced a total of 60 Coriell DNA samples (One Coriell DNA sample at 12 dilutions and additional 48 Coriell DNA samples with different genotypes) and 20 oral swabs samples from healthy individuals. We used two types of flowcells, a smaller MinION Flongle flow cell with up to 2.8Gb output, and a regular Spot On R9.4.1 flow cell with 20Gb output (**Supplementary table 2**). The library was prepared using the SQK-LSK110 Ligation Sequencing Kit (Oxford Nanopore Technologies). Samples were barcoded using a Nanopore PCR barcoding expansion (EXP-PBC096 PCR Barcoding Expansion). For each library, we took approximately 100ng of the PCR purified product from first round of PCR. The barcoding mix was prepared by adding 1μl of 10uM PCR barcode to 25μl LongAmp Taq 2x master mix (NEB) and 100ng of first-round PCR product. The volume was adjusted to 50μl, and barcoding was performed at 12 rounds of PCR cycles. The purified libraries were pooled to a total 1μg followed by DNA repair and end-prep using NEBNext FFPE DNA Repair Mix and NEBNext Ultra II End repair / dA-tailing Module reagents in accordance with manufacturer’s instructions. Adaptor ligation was performed using Adapter Mix F (AMX-F) and Quick T4 Ligase. For targeted sequencing, we used 250 μl Short Fragment Buffer (SFB) for the final wash to retain shorter amplicons. The final library was eluted on in 15μl elution buffer (EB). We loaded the maximum recommended quantities of libraires in all six runs (20fm libraries on a Flongle flow cell and 50fm of a Spot flow cell). The samples were sequenced on a MinION Mk1C sequencer.

**Table E1**: Anchored primers for 8-plex PCR

| **New name** | **Final primer conc.** | **SNP position** | **SNP** | **Sequence** | **Product length (bp)** | **Tm** |
| --- | --- | --- | --- | --- | --- | --- |
| SLCO1B1-exon-388-FP | 0.3 µM | rs2306283 | 388A>G | TTTCTGTTGGTGCTGATATTGCCTTGACCAAGATATAACCACTCCT | 675 | 64.9 |
| SLCO1B1-exon-388-RP | 0.3 µM |  |  | ACTTGCCTGTCGCTCTATCTTCGGTTTATCATCCAGTTCAGATGGA |  | 66.4 |
| SLCO1B1-exon-521-FP | 0.3 µM | rs4149056 | 521T>C | TTTCTGTTGGTGCTGATATTGCCATGAGGAACTATGAGTCCATTAG | 605 | 63.5 |
| SLCO1B1-exon-521-RP | 0.3 µM |  |  | ACTTGCCTGTCGCTCTATCTTCCAAGAATGCATGGTTCTTATTCAC |  | 63.8 |
| CYP2E1-RsaI_PstI_FP | 0.3 µM | rs2031920, rs3813867 | −1053C>T, −1293G>C | TTTCTGTTGGTGCTGATATTGCCAGTATTATGGCCAGAGGACT | 1047 | 64.2 |
| CYP2E1-RsaI_PstI_RP | 0.3 µM |  |  | ACTTGCCTGTCGCTCTATCTTCGACACAATAGAGCTCCACATTG |  | 64.4 |
| CYP2E1-DraI_FP | 0.3 µM | rs6413432 | T7632A | TTTCTGTTGGTGCTGATATTGCGCTAATGGTCACTTGTGGTCT | 921 | 65.5 |
| CYP2E1-DraI_RP | 0.3 µM |  |  | ACTTGCCTGTCGCTCTATCTTCCTAGTGCAGAGTAGTCGAAAGTT |  | 65.1 |
| NAT2-FP | 0.3 µM | rs1801279, rs1041983, rs1801280, rs1799929, rs1799930, rs1208, rs1799931 | 191G>A, 282C>T, 341T>C, 481C>T, 590G>A, 803A>G, 857G>A | TTTCTGTTGGTGCTGATATTGCCATGGAGTTGGGCTTAGAGG | 820 | 65.4 |
| NAT2-RP | 0.3 µM |  |  | ACTTGCCTGTCGCTCTATCTTCGAGTTGGGTGATACATACACAAGG |  | 66.2 |
| CYP3A5-FP | 0.3 µM | rs776746 | 6986A>G | TTTCTGTTGGTGCTGATATTGCGTCCTTGTGAGCACTTGATG | 551 | 64.3 |
| CYP3A5-RP | 0.3 µM |  |  | ACTTGCCTGTCGCTCTATCTTCCTTTCACTAGCACTGTTCTGATC |  | 64.3 |
| AADAC-FP | 0.3 µM | rs1803155 | G>A | TTTCTGTTGGTGCTGATATTGCCTTGCTTCTCAGCTCCTTG | 485 | 63.7 |
| AADAC-RP | 0.3 µM |  |  | ACTTGCCTGTCGCTCTATCTTCGAGATCATATTGACAGGTGATGAC |  | 64 |
| SLCO1B-Intron FP | 0.48 µM | rs4149032 | C>T | TTTCTGTTGGTGCTGATATTGCGGATGAATTGTACAATCATTCCTTAG | 1232 | 63.2 |
| SLCO1B-Intron RP | 0.48 µM |  |  | ACTTGCCTGTCGCTCTATCTTCTACCGTCTCCAGCTATATGTTC |  | 64.2 |

**Supplementary Table E2**: Sequencing statistics for 180 (Coriell DNA, oral swabs, DNA from whole blood) samples analyzed on Flongle and Spot On R.9.4.1 flowcell

| **Sample type** | **Total (n)** | **Amount of library (ng)/sample** | **Flow cell type** | **Run time (hr)** | **Active pores (n)** | **Median Reads passed/sample** | **Median yield (Mb)/sample** | **Median quality score** | **Median depth (X)** |
| --- | --- | --- | --- | --- | --- | --- | --- | --- | --- |
| Batch 1 | 19 | 52.6 | Flongle | 24 | 65 | 19,864 | 15.8 | 12.8 | 992 |
| Batch 2 | 41 | 23.8 | Spot On (R9.4.1) | 48 | 1096 | 86,184 | 61.6 | 13.8 | 3540.5 |
| Batch 3 | 20 | 33.3 | Spot On (R9.4.1) | 48 | 1208 | 122,457 | 90.3 | 13.5 | 12,143 |
| Batch 4 | 40 | 25 | Spot On (R9.4.1) | 48 | 1354 | 100,000 | 76.9 | 13.8 | 4604.5 |
| Batch 5 | 38 | 25 | Spot On (R9.4.1) | 24 | 557 | 50,000 | 35.5 | 14.1 | 2168.5 |
| Batch 6 | 22 | 40 | Spot On (R9.4.1) | 24 | 333 | 46,940 | 36.2 | 12.3 | 2670.7 |

**Table E3. Final population parameter estimates for rifampicin**

| **Parameter description** | **Typical value (95% CI) ^c^** |
| --- | --- |
| Bioavailability | 1 Fixed |
| Absorption Lag Time (h) | 0.820 (0.722 – 0.918) |
| Absorption rate constant (1/h) | 1.30 (1.11 – 149) |
| Volume of distribution (L) ^a^ | 55.3 (52.3 – 58.3) |
| Clearance (L/h) ^a^ |  |
| HIV-1-uninfected patients | 24.7 (20.8 – 28.6) |
| HIV-1-infected patients on LPV/r | 13.4 (9.26 – 19.5) |
| HIV-1-infected patients not on LPV/r | 21.7 (18.4 – 25.5) |
| Effect of AADAC intron (rs1803155) on clearance |  |
| Heterozygote and Wild type | 1 (Reference) |
| Homozygous mutant (%) ^b^ | -17.3% (-29.9 – -2.50%) |
| **Parameter variability (CV%)** |  |
| BSV Clearance | 38.3 (31.5 – 45.4) |
| BOV Bioavailability | 28.6 (23.0 –34.2) |
| BOV Lag Time | 69.8 (56.5 – 84.5) |
| BOV Absorption rate constant | 63.2 (50.4 – 77.2) |
| **Residual unexplained variability** |  |
| Additive error (mg/L) | 0.170 (0.137 – 0.203) |
| Proportional error (%) | 16.0 (14.3 – 17.7) |
| Abbreviations: %CV, coefficient of variation; BOV, between-occasion variability; BSV, between-subject variability.  ^a^ Allometrically scaled with fat-free mass (median 43.2 kg)  ^b^ Compared to heterozygote and wild type  c Calculated by $CV\%= \sqrt{e^{\omega^{2}}-1}\cdot100\%$, where ω^2^ represents the variance | |

**Table E4. Final population parameter estimates for isoniazid**

| **Parameter description** | **Typical value (95% CI) ^b^** |
| --- | --- |
| Bioavailability | 1 Fixed |
| Absorption mean transit time (h) | 0.450 (0.324 – 0.577) |
| Number of transit compartments (n) | 2.56 (2.29 – 2.83) |
| Absorption rate constant (1/h) | 1.05 (0.954 – 1.15) |
| Volume of distribution (L) ^a^ | 20.0 (15.7 – 24.3) |
| Clearance (L/h) ^a^ |  |
| Slow acetylators | 11.4 (10.0 – 12.9) |
| Intermediate acetylators | 28.2 (24.0 – 33.2) |
| Rapid acetylators | 47.5 (38.3 – 58.9) |
| Peripheral volume of distribution (L) | 25.6 (22.7 – 28.4) |
| Intercompartmental clearance (L/h) | 17.5 (13.6 – 21.4) |
| **Parameter variability (CV%)** |  |
| BSV Clearance | 35.0 (29.1 – 41.1) |
| BOV Bioavailability | 30.7 (25.7 – 35.8) |
| BOV Absorption mean transit time | 106 (67.8 – 160) |
| BOV Absorption rate constant | 15.1 (7.76 – 22.5) |
| **Residual unexplained variability** |  |
| Additive error (mg/L) | 0.00250 (0 – 0.0.0276) |
| Proportional error (%) | 13.0 (11.9 – 14.1) |
| Abbreviations: %CV, coefficient of variation; BOV, between-occasion variability; BSV, between-subject variability.  ^a^ Allometrically scaled with fat-free mass (median 43.2 kg)  ^b^ Calculated by $CV\%= \sqrt{e^{\omega^{2}}-1}\cdot100\%$, where ω^2^ represents the variance | |
